# Identification and validation of heterotypic cell-in-cell structure as an adverse prognostic predictor for young patients of resectable pancreatic ductal adenocarcinoma

**DOI:** 10.1101/2020.07.08.20148825

**Authors:** Hongyan Huang, Meifang He, Yanbin Zhang, Bo Zhang, Zubiao Niu, You Zheng, Wen Li, Peilin Cui, Xiaoning Wang, Qiang Sun

## Abstract

**OBJECTIVES:** A proportion of resectable pancreatic ductal adenocarcinoma (PDAC) patients display poorer survival due to profound local immune suppression. However, a pathological/morphological parameter that could functionally read out immune evasion and predict patient survival has not been defined. This study investigated the feasibility of heterotypic cell-in-cell (CIC) structures for immune cell cannibalism by tumor cells to serve as a parameter for survival prediction in resectable PDAC patients.

**METHODS:** A total of 410 samples from PDAC patients were examined using the methods of “EML” multiplex staining or immunohistochemistry (IHC). Prognostic CIC candidates were initially identified in samples plotted in tissue microarray (n=300), then independently validated in specimens from the First Affiliated Hospital of Sun Yat-Sen University (n=110). The Kaplan–Meier estimator and/or the Cox regression model were used for univariate and multivariate analysis. A nomogram was made using the Regression Modeling Strategies.

**RESULTS:** CICs were prevalent in cancerous (203/235) but not non-malignant tissues (15/147). Among the 4 CIC subtypes identified, 2 heterotypic subtypes with tumor cells internalizing CD45^+^ lymphocytes (LiT, mOS = 8 vs. 14.5 months, *p* = 0.008) or CD68^+^ monocytes (MiT, mOS = 7.5 vs. 15 months, *p* = 0.001), and overall CICs (oCIC, mOS = 10 vs. 27 months, *p* = 0.021), but not homotypic CICs (TiT, *p* = 0.089), were identified in univariate analysis as adverse prognostic factors of overall survival (OS) of PDAC. Notably, through cannibalism of immune cells by tumor cells, heterotypic CICs (L/MiT: LiT plus MiT) could independently predict shorter OS (HR = 1.85, *p* = 0.008) in multivariate analysis, with a performance comparable or even superior to traditional clinicopathological parameters such as histological grade (HR = 1.78, *p* = 0.012) and TNM stage (HR=1.64, *p* = 0.108). This was confirmed in the validation cohort, where L/MiT (HR = 1.71, *p* = 0.02) and tumor–node–metastasis (TNM) stage (HR = 1.66, *p* = 0.04) were shown to be independent adverse prognostic factors. Moreover, L/MiT stood out as the most prominent contributor in nomogram models constructed for survival prediction (area under the curve = 0.696 at 14 months), the dropout of which compromised prediction performance (area under the curve = 0.661 at 14 months). Furthermore, stratification analysis indicated that L/MiT tended preferentially to impact young and female patients (HR = 11.61, *p* < 0.0001, and HR = 9.55, *p* = 0.0008, respectively) in particular with early-stage and low-grade PDAC (HR = 2.37, *p* < 0.0001, and HR = 2.19, *p* < 0.0001, respectively), while TNM stage demonstrated little preference.

**CONCLUSION:** This was the first CIC profiling to be performed in PDAC, and is currently largest for human tumors. Subtyped CICs, as a valuable input to the traditional variables such as TNM stage, represent a novel type of prognostic factor. The formation of heterotypic L/MiT may be a surrogate for local immune evasion and predict poor survival, particularly in young female patients of resectable PDAC.

**Study Highlights:** *Prior knowledge:*

- The post-operation survival periods of resectable pancreatic ductal adenocarcinoma (PDAC) patients range widely, and the search for reliable prognostic biomarkers is warranted.
- Although profound local immune suppression is implicated in PDAC progression and poor patient survival, a prognostic marker to read immune evasion in situ is not yet available.
- The impact of subtyped cell-in-cell (CIC) structures, which target either tumor or immune cells for internalization and death, on PDAC patient survival is not clear.

*Novelty of study:*

- This study presents the first CIC subtype profiling in PDAC, which is currently the largest of its type for human cancers.
- Subtyped CIC structures were identified and confirmed independently as a valuable prognostic factor for PDAC patients, with a performance comparable or superior to traditional variables such as tumor–node–metastasis (TNM) stage.
- The L/MiT heterotypic CIC subtype, surrogating a type of cellular immune evasion, could independently predict poor survival, particularly for young female patients of resectable PDAC.

## Introduction

Pancreatic ductal adenocarcinoma (PDAC) has a poor prognosis, with an overall 5-year survival rate of less than 10%, and a median survival time of less than 6 months if left untreated ^1^. The incidence of pancreatic cancer is higher in males than in females and related to smoking history. Surgical resection remains the only potentially curative treatment for pancreatic cancer and the addition of adjuvant chemotherapy has been shown to improve survival rates. There have been some optimistic results demonstrating a further improvement in survival with the administration of chemo-radiotherapy in the neo-adjuvant setting for the patients with local advanced disease ^2^. Between 20% and 30% of PDAC cases are resectable at diagnosis; however, patients’ post-operative survival periods vary widely, irrespective of active therapeutic interventions ^3^. Therefore, extensive efforts have been made to identify biomarkers that may identify patients with an improved prognosis ^4, 5^. Although serological markers such as carbohydrate antigen 19-9 (CA19–9) and carcino-embryonic antigen (CEA) have demonstrated a predictive performance in some models ^5-7^, inconsistencies have been reported, and the inherent indirectness of serological markers still exists. This highlights the need to identify novel biomarkers, especially pathological/morphological markers that can directly and functionally predict malignancy.

The poor prognosis of PDAC was believed to be related to comprehensive local immunosuppression within tumor tissues, where complex interactions between tumor, stroma, and immune cells establish an immunosuppressive tumor microenvironment (TME), which promotes cancer growth and progression ^8^. This idea is supported by the fact that few immunotherapy approaches have succeeded with PDAC, although promising therapeutic efficacies have been demonstrated for other types of solid tumor ^3, 9^. Also, enrichment for M2 macrophages was shown to be associated with patient survival by PDAC stratification analysis ^10^. Recent studies have indicated that, in addition to immunosuppressive factors such as cytokines and cell surface proteins, the interaction between tumor and immune cells could give rise to the so-called cell-in-cell structures (CICs) that might serve as a novel mechanism of immune evasion ^11-14^.

“CICs” refers to the presence of one or more cells inside a host cell, which generally leads to the death of inner cells ^15^. CICs are prevalent in a wide range of human tumors and have different subtypes, resulting from active interactions between certain types of TME cells ^16^, including homotypic CICs formed between tumor cells, and heterotypic CICs formed via internalization of mesenchymal stem cells or immune cells into tumor cells ^17-20^. Cell death via different types of CICs has been found to give rise to multiple mechanisms, such as the homotypic CIC-mediated entosis ^21^, heterotypic CIC-mediated cannibalism ^22^, and emperitosis ^23^. Therefore, CICs are ideal candidates for the functional readout of malignant progression. CICs have been identified in several types of cancers ^24^, and have been suggested to be related to either tumor suppression ^25^ or progression ^26, 27^. However, due to the lack of systematic analyses associating the presence of distinct types of CICs with patient clinicopathological history, their impact on patients’ prognosis remains to be elucidated. On these grounds, the contribution of different CICs, especially heterotypic CICs, to patient prognosis has yet to be established. The aim of this study was to explore the feasibility of using subtyped CICs as a kind of functional biomarker that can read out tumor malignancy and predict patient survival in PDAC.

## Materials and methods

### Human tumor tissue microarray and PDAC tissue

Two human tumor tissue microarrays (TMA) with paired samples of resectable cancer and non-malignant pancreas tissues from 153 PDAC patients were purchased from Shanghai Outdo Biotech Co. Ltd (HPan-Ade180Sur-01 and HPan-Ade120Sur-01). The Outdo Biotech is a leading company in human/animal tissue microarrays (TMA) and “clinical-type” gene chips (CTGCs) in China. All tissues were collected under the highest ethical standards with the donors being fully informed and their consent being obtained. The samples were collected from patients between 2004 and 2008 and stored and transported at −80°C. The cases were routinely followed up by professional doctors. The TMA slide was prepared from formalin-fixed, paraffin-embedded cancer and paired non-malignant tissue. In total, there were 300 cores on 2 slides, including 153 cases of pancreatic cancer tissues and 147 cases of non-malignant tissues (not plotted for 6 patients). The diameter of each core was 1.5 mm (1.76 mm^2^/core). For validation, tissue sections were collected from 110 resectable PDAC patients who had received surgery at the Department of Hepatobiliary Surgery, the First Affiliated Hospital of Sun Yat-sen University (Guangzhou, China), with the institutional research ethics committee reviewing and approving the research. All of the patients received ultrasound and computed tomography scans prior to surgery and none received systemic chemotherapy or radiotherapy preoperatively. All specimens were diagnosed by pathological examination after surgery.

### TMA and tissue specimen staining, and antibodies

The “EML method”, a multiplexing method based on the technique of tyramide signal amplification (TSA) ^16^, was employed to subtype CICs. Through this method, tissues were simultaneously stained with antibodies against E-cadherin for epithelial cancer cells, CD45 for leukocytes, and CD68 for macrophages. Slides were routinely de-paraffinized with the xylene-ethanol method and baked at 65°C for 1.5 hours. Antigen retrieval was performed in citrate acid buffer by microwaving for 15 minutes after boiling, followed by 1 hour blocking in 5% bovine serum albumin (BSA) made in Tris-buffered saline (TBS). Samples were first stained with anti-CD45 antibody (mouse mAb from Boster, BM0091) at a dilution of 1:400 using Opal Multiplex tissue staining kit (Perkin Elmer, NEL791001KT) according to the manufacturer’s standard protocol. CD45 molecules were subsequently labeled with Cyanine 5 fluorophore. The slides were then incubated with mixed antibodies for E-Cadherin (1:200, mouse mAb from BD Biosciences, 610181) and CD68 (1:200, rabbit pAb from Proteintech, 25747–1-AP), followed by Alexa Fluor 568 secondary anti-rabbit antibody (Invitrogen, A11036) and Alexa Fluor 488 anti-mouse antibody (Invitrogen, A11029). Samples were also labeled with single fluorophore to acquire spectral signatures. All slides were counterstained with DAPI to show nuclei, before being mounted with Antifade reagent (Invitrogen, Carlsbad, CA, USA) and cover slips, and then sealed with clear nail polish. For validation, tissue sections were stained by hematoxylin and eosin (H&E) and immunohistochemistry (IHC) with each of the antibodies indicated above, following the protocol provided by Cell Signaling Technology (https://www.cellsignal.com/contents/resources-protocols/immunohistochemistry-protocol-paraffin-for-signalstain-boost-detection-reagent/ihc-paraffin-signalstain).

### Multispectral imaging and analysis

Multispectral images were taken with TMA modules of Vectra® Automated Imaging System (Perkin Elmer) by a 20x objective lens (Figure S1A-C). A nuance system (Perkin Elmer) was used to build libraries of each spectrum (DAPI, 488, 568, and Cy5-650) and unmix multispectral images with high contrast and accuracy (Figure 1A-D). inForm automated image analysis software package (Perkin Elmer) was used for batch analysis of multispectral images based on specified algorithms.

**Figure 1.**
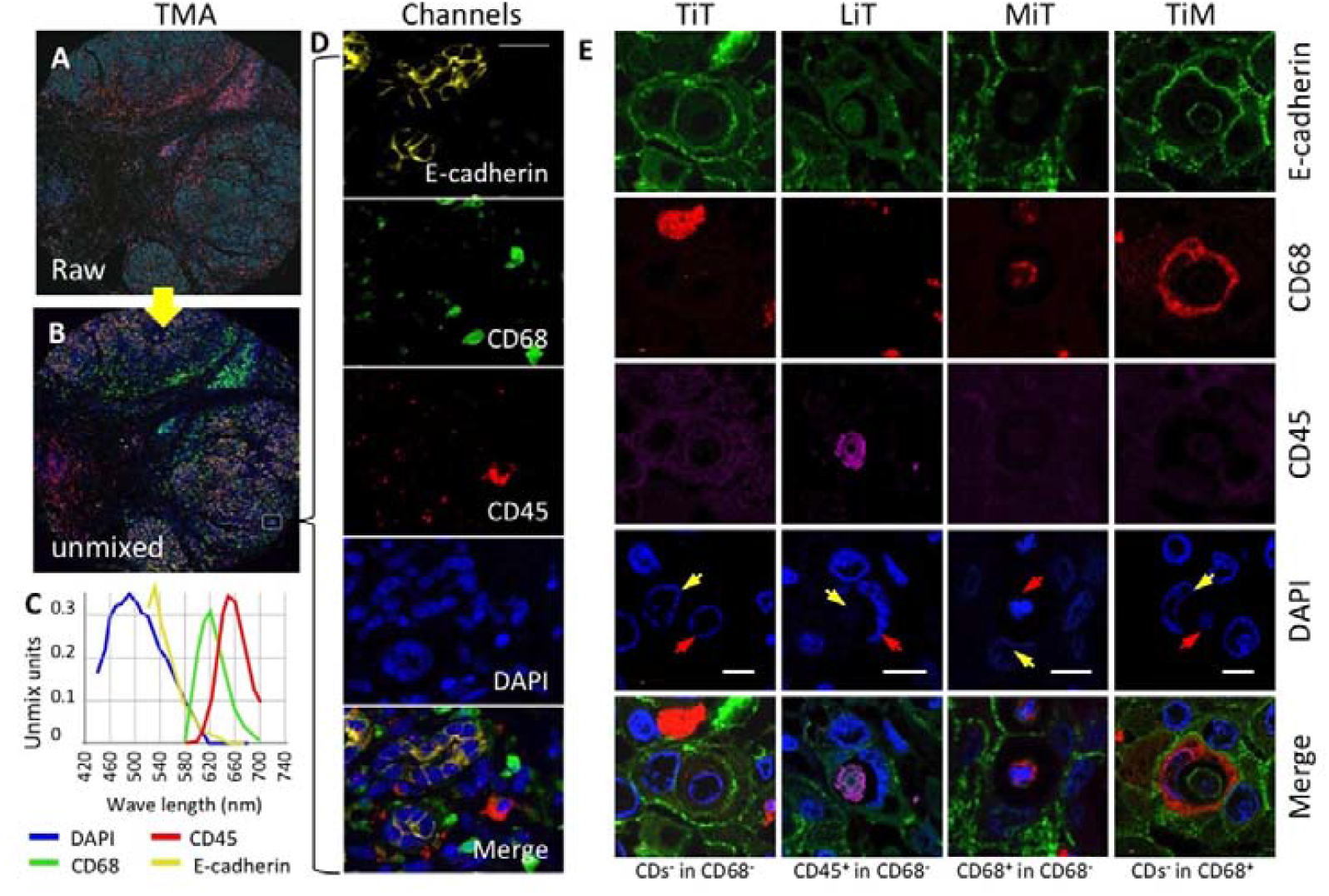
Identification of 4 subtypes of CIC structures in PDAC. (**A**) Unprocessed composite core multiplex stained with E-cadherin, CD68, CD45, and DAPI. (**B**) Unmixed composite core pseudo-colored with yellow for E-cadherin, green for CD68, red for CD45, and blue for DAPI. (**C**) Spectral parameters for image unmixing. Fluorescent signals for different targets were captured based on the spectra indicated. (**D**) Images of single or merged channels displaying the boxed region in (**C**). Scale bar: 20 μm. (**E**) Representative images for the 4 CIC subtypes as indicated; scale bars: 5 μm;

### CIC profiling and quantification

Cellular structures were scored as CICs where one or more cells morphologically were fully enclosed within another cell with a crescent nucleus. As CICs can result in inner cell death, we scored all structures displaying CIC morphology irrespective of whether inner cells were dead or live. Cell boundaries were identified by E-cadherin, which labels cell membranes, and/or CD68, which labels cell bodies. CIC subtypes were defined based on the types of cells involved: TiT for E-cadherin^+^ cells inside E-cadherin^+^ cells; TiM for E-cadherin^+^ cells inside CD68^+^ cells, MiT for CD68^+^ cells inside E-cadherin^+^ cells, LiT for CD45^+^ cells inside E-cadherin^+^ cells. For efficient quantification in TMA, CICs were usually first screened in a composite image of 4 fluorescent channels and then confirmed in unmixed channels. For quantification in validation specimens, subtyped CICs were counted based on IHC staining with reference to H&E staining (Figure S2). Double-blind reviews were performed for all the CIC quantifications.

### Statistical analysis

Statistical analysis was performed using the SPSS 20.0 (IBM Corp., NY, USA) and EmpowerStats (http://www.empowerstats.com/) software systems, which wraps R software. Study data were collected on standard forms and checked for completeness. All data were described using median (min–max) for continuous variables like follow-up times, while frequencies (percent) were used for categorical variables. Overall survival (OS) was defined as time from the date of surgery to death or to the most recent contact or visit. Survival times were analyzed by the Kaplan–Meier method, and the differences in survival times were compared by the log-rank test. Univariate and multivariate survival analyses were performed using the Cox proportional-hazard models, and hazard ratios (HRs) (95% confidence interval) were calculated. The association between clinicopathological factors and the number of CICs was analyzed using the Chi-square test or Fisher’s exact test. The nomogram was formulated based on the results of multivariate logistic regression analysis by Regression Modeling Strategies, which proportionally converts each regression coefficient in multivariate logistic regression to a 0-to-100-point scale as described elsewhere ^28^. The area under the curve (AUC) calculation was performed and graphed with EmpowerStats software. For all analyses, a two-sided *p* value of less than 0.05 was considered statistically significant.

## Results

### Patient characteristics

In total, 410 specimens from 147 pair-matched cancer and non-malignant tissues and 116 cancer-only tissues, were included in this study. Out of these, 300 specimens, consisting of 147 case-matched non-malignant pancreas controls and 153 cancer tissues (note: only 125 cancer tissues were included into the final analysis due to missing information or tissue detachment during staining), were plotted on TMA and used as the discovery cohort. The validation cohort consisted of additional 110 specimens collected from the First Affiliated Hospital of Sun Yat-sen University. As listed in Table 1, the clinical characteristics of the 235 patients included in the final analysis were generally comparable between the discovery and validation cohorts. Most of the patients were male (60%) and the median age of patients was 62 (27–78) years old. The majority of the PDAC cases occurred in the pancreas head (72%) and were diagnosed at an early stage (75% at TNM I+II; 69% at high and moderate differentiation), with only small proportion of patients having experienced distant metastasis (11%). All of the patients had follow-ups until their death or until their most recent contact or visit (November 30, 2014 for the discovery cohort, and March 30, 2011 for the validation cohort). At the time of their most recent contact, 72 of the 235 patients were still alive, and 163 patients had succumbed to their disease. Median follow-up time was 12.0 months (0.2–87.0 months) and 30.5 months (0.1–57.8 months) in the discovery cohort and validation cohort, respectively.

**Table 1.**
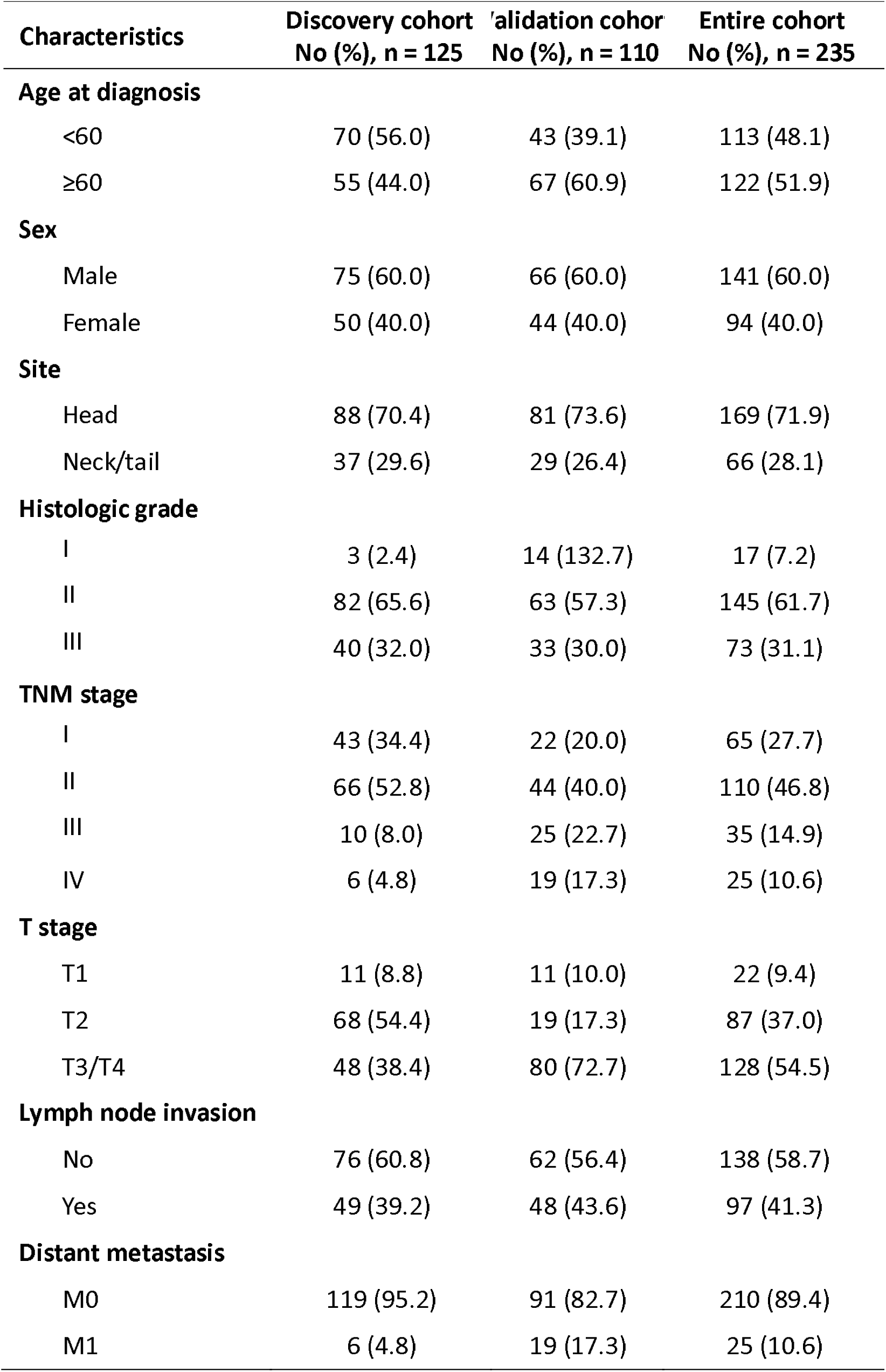
Characteristics for different cohorts of patients.

### CIC profiling in PDAC tissues

The CICs in the discovery cohort of specimens were quantified as described in our previous study ^16^, and only the structures with inner cells fully enclosed were counted (Figure S1B). CICs were minimally detected in non-malignant tissues (15/147) but were prevalent in cancer tissues (97/125) (*p* < 0.0001) (Figure S1D-E). Four CIC subtypes were identified (Figure 1E): tumor cells inside tumor cells (TiT), lymphocytes (CD45^+^) inside tumor cells (CDs^-^) (LiT), tumor cells inside <u>m</u>acrophages (TiM), and <u>m</u>acrophages inside tumor cells (MiT). The subtype of TiT constituted the majority (70.1%) of overall CICs (oCICs) (Figure 2A-B). Out of the 3 heterotypic CICs, LiT accounted for 7.3%. The remaining 2 CIC subtypes were formed between tumor cells and macrophages (CD68^+^), which mutually engulfed each other, generating MiT (19.3%) and TiM (3.6%) (Figure 2A-C). Identity analysis indicated that tumor cells were the major engulfer (96.4%) over macrophages (3.6%) (Figure 2D), and that both tumor cells (73.4%) and immune cells (7.3% CD45^+^ and 19.3% CD68^+^) could be internalized as inner cells (Figure 2E). A similar CIC profile was identified in the validation cohort (Figure 2F-J).

**Figure 2.**
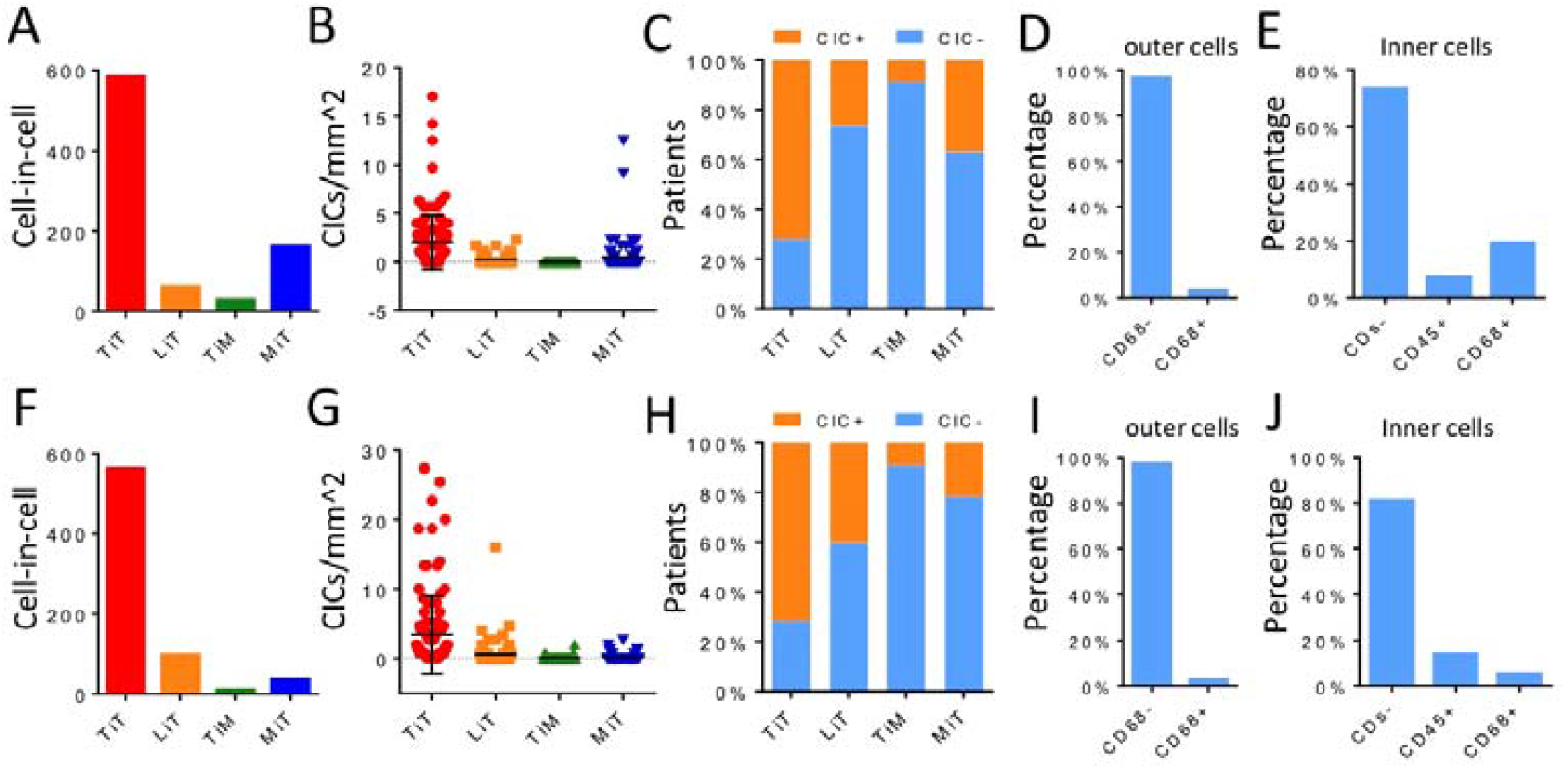
CIC profiling in discovery (A–E) and validation (F–J) cohorts. Profiles of CICs subtypes depicted for all CIC counts (**A, F**), for CIC counts normalized by core area (**B, G**), for patient percentages (**C, H**), for outer cell identities (**D, I**), and for inner cell identities (**E, J**).

### Association of CICs with clinicopathological characteristics

In the discovery cohort, while the presence of oCICs was not significantly associated with the known clinicopathological characteristics, specific CIC subtypes did show associations (Table S1a). Homotypic TiT was more frequently detected in tissues of older patients (45/70 for < 60 years old vs. 45/55 for ≥ 60 years old, *p* = 0.03); however, this was not confirmed in the validation dataset. Heterotypic MiT and L/MiT (for LiT plus MiT) tended to be more frequently present in tumor tissues of a late TNM stage (35/109 vs. 11/16, *p* = 0.005) and low tumor differentiation (37/85 vs. 25/40, *p* = 0.048), respectively, which were confirmed in the validation cohort (Table S1b). When combining the discovery and validation datasets, the presence of homotypic TiT also demonstrated a significant association with late TNM stage (p = 0.0032) (Table S1c). These results are consistent with the notion that later stage tumor cells more frequently engage in cannibalistic activities.

### The presence of heterotypic CICs is associated with shorter postoperative survival

Univariate analysis revealed that tumor grade, TNM stage, lymph node (LN) invasion, and distant metastasis were significantly associated with postoperative survival in the discovery cohort, the validation cohort, and the combined cohort (Table 2 and S2). Of these traditional variables, TNM stage was likely, and expectedly, the most consistent and representative survival classifier (median overall survival time (mOS): 7 vs. 14 months; *p* < 0.0001) across the 3 cohorts (Table 2 and Figure S3). Notably, the presence of oCICs was associated with a shorter OS with a better performance but bigger variation (mOS: 10 vs. 27 months; *p* = 0.021) (Table 2 and Figure 3), which could be attributed to the different contributions of CIC subtypes as revealed by subtype analysis. As shown in Table 2 and Figure 3, while homotypic TiT was a weak classifier (mOS: 11 vs. 21 months; *p* = 0.089), heterotypic L/MiT displayed a stronger prognostic power in predicting shorter postoperative survival (mOS: 8 vs. 31 months; *p* < 0.0001), which was validated in the patients from The First Affiliated Hospital of Sun Yat-sen University. The adverse prognostic role of heterotypic CICs may have been specific for CICs of the tumor cell engulfer (L/MiT) as the presence of TiM, the heterotypic CIC subtype of macrophage engulfer, did not significantly impact patient survival (mOS: 7 vs. 13 months, *p* = 0.184) (Table 2, S2, and Figure S3).

**Table 2.**
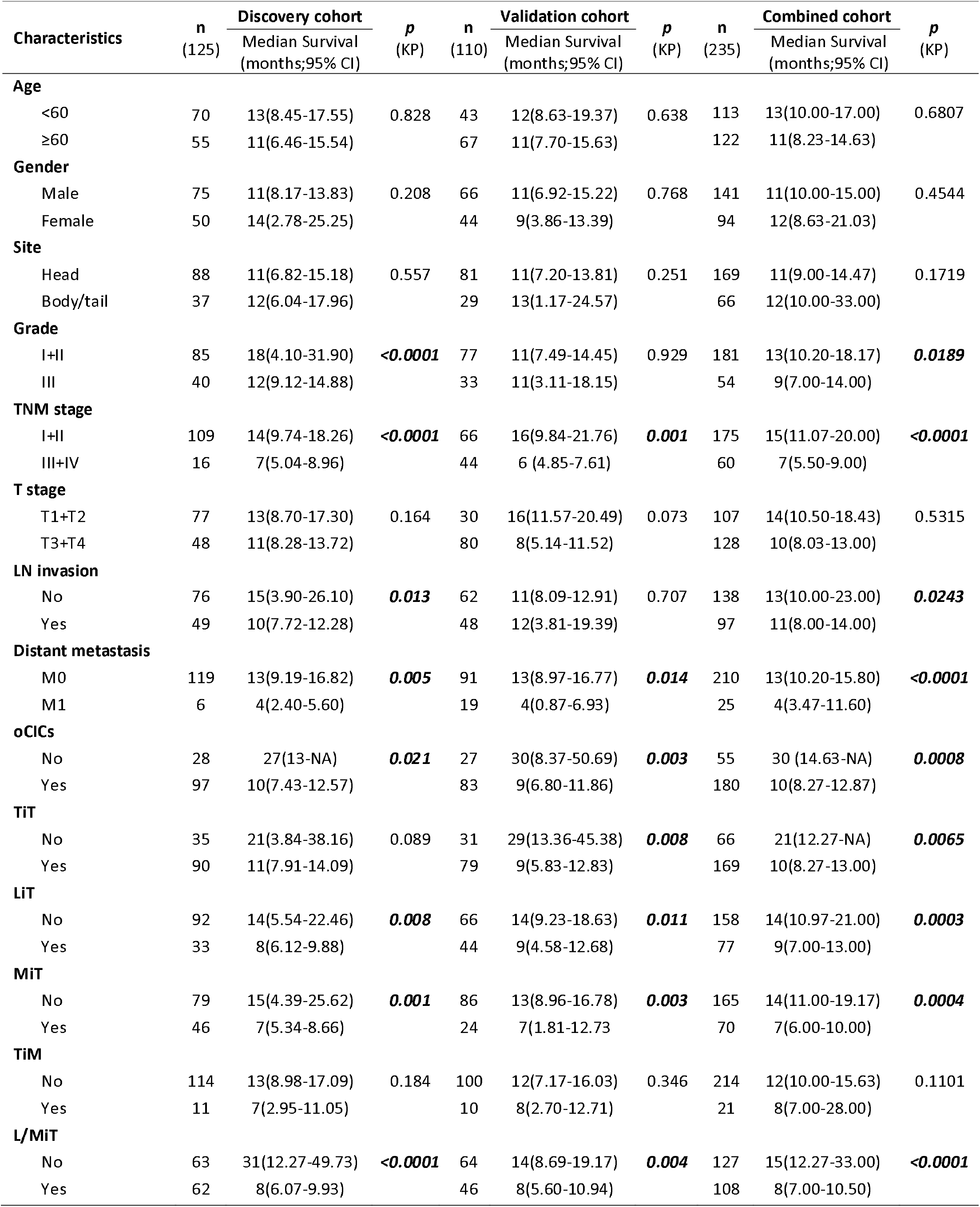
The univariate Kaplan–Meier survival analysis for PDAC patients.

| Characteristics | n<br>(125) | Discovery cohort<br>Median Survival<br>(months;95% CI) | p<br>(KP) | n<br>(110) | Validation cohort<br>Median Survival<br>(months;95% CI) | p<br>(KP) | n<br>(235) | Combined cohort<br>Median Survival<br>(months;95% CI) | p<br>(KP) |
| --- | --- | --- | --- | --- | --- | --- | --- | --- | --- |
| **Age** |  |  |  |  |  |  |  |  |  |
| <60 | 70 | 13(8.45-17.55) | 0.828 | 43 | 12(8.63-19.37) | 0.638 | 113 | 13(10.00-17.00) | 0.6807 |
| ≥60 | 55 | 11(6.46-15.54) |  | 67 | 11(7.70-15.63) |  | 122 | 11(8.23-14.63) |  |
| **Gender** |  |  |  |  |  |  |  |  |  |
| Male | 75 | 11(8.17-13.83) | 0.208 | 66 | 11(6.92-15.22) | 0.768 | 141 | 11(10.00-15.00) | 0.4544 |
| Female | 50 | 14(2.78-25.25) |  | 44 | 9(3.86-13.39) |  | 94 | 12(8.63-21.03) |  |
| **Site** |  |  |  |  |  |  |  |  |  |
| Head | 88 | 11(6.82-15.18) | 0.557 | 81 | 11(7.20-13.81) | 0.251 | 169 | 11(9.00-14.47) | 0.1719 |
| Body/tail | 37 | 12(6.04-17.96) |  | 29 | 13(1.17-24.57) |  | 66 | 12(10.00-33.00) |  |
| **Grade** |  |  |  |  |  |  |  |  |  |
| I+II | 85 | 18(4.10-31.90) | **<0.0001** | 77 | 11(7.49-14.45) | 0.929 | 181 | 13(10.20-18.17) | **0.0189** |
| III | 40 | 12(9.12-14.88) |  | 33 | 11(3.11-18.15) |  | 54 | 9(7.00-14.00) |  |
| **TNM stage** |  |  |  |  |  |  |  |  |  |
| I+II | 109 | 14(9.74-18.26) | **<0.0001** | 66 | 16(9.84-21.76) | **0.001** | 175 | 15(11.07-20.00) | **<0.0001** |
| III+IV | 16 | 7(5.04-8.96) |  | 44 | 6 (4.85-7.61) |  | 60 | 7(5.50-9.00) |  |
| **T stage** |  |  |  |  |  |  |  |  |  |
| T1+T2 | 77 | 13(8.70-17.30) | 0.164 | 30 | 16(11.57-20.49) | 0.073 | 107 | 14(10.50-18.43) | 0.5315 |
| T3+T4 | 48 | 11(8.28-13.72) |  | 80 | 8(5.14-11.52) |  | 128 | 10(8.03-13.00) |  |
| **LN invasion** |  |  |  |  |  |  |  |  |  |
| No | 76 | 15(3.90-26.10) | **0.013** | 62 | 11(8.09-12.91) | 0.707 | 138 | 13(10.00-23.00) | **0.0243** |
| Yes | 49 | 10(7.72-12.28) |  | 48 | 12(3.81-19.39) |  | 97 | 11(8.00-14.00) |  |
| **Distant metastasis** |  |  |  |  |  |  |  |  |  |
| M0 | 119 | 13(9.19-16.82) | **0.005** | 91 | 13(8.97-16.77) | **0.014** | 210 | 13(10.20-15.80) | **<0.0001** |
| M1 | 6 | 4(2.40-5.60) |  | 19 | 4(0.87-6.93) |  | 25 | 4(3.47-11.60) |  |
| **oCICs** |  |  |  |  |  |  |  |  |  |
| No | 28 | 27(13-NA) | **0.021** | 27 | 30(8.37-50.69) | **0.003** | 55 | 30 (14.63-NA) | **0.0008** |
| Yes | 97 | 10(7.43-12.57) |  | 83 | 9(6.80-11.86) |  | 180 | 10(8.27-12.87) |  |
| **TiT** |  |  |  |  |  |  |  |  |  |
| No | 35 | 21(3.84-38.16) | 0.089 | 31 | 29(13.36-45.38) | **0.008** | 66 | 21(12.27-NA) | **0.0065** |
| Yes | 90 | 11(7.91-14.09) |  | 79 | 9(5.83-12.83) |  | 169 | 10(8.27-13.00) |  |
| **LiT** |  |  |  |  |  |  |  |  |  |
| No | 92 | 14(5.54-22.46) | **0.008** | 66 | 14(9.23-18.63) | **0.011** | 158 | 14(10.97-21.00) | **0.0003** |
| Yes | 33 | 8(6.12-9.88) |  | 44 | 9(4.58-12.68) |  | 77 | 9(7.00-13.00) |  |
| **MiT** |  |  |  |  |  |  |  |  |  |
| No | 79 | 15(4.39-25.62) | **0.001** | 86 | 13(8.96-16.78) | **0.003** | 165 | 14(11.00-19.17) | **0.0004** |
| Yes | 46 | 7(5.34-8.66) |  | 24 | 7(1.81-12.73) |  | 70 | 7(6.00-10.00) |  |
| **TiM** |  |  |  |  |  |  |  |  |  |
| No | 114 | 13(8.98-17.09) | 0.184 | 100 | 12(7.17-16.03) | 0.346 | 214 | 12(10.00-15.63) | 0.1101 |
| Yes | 11 | 7(2.95-11.05) |  | 10 | 8(2.70-12.71) |  | 21 | 8(7.00-28.00) |  |
| **L/MiT** |  |  |  |  |  |  |  |  |  |
| No | 63 | 31(12.27-49.73) | **<0.0001** | 64 | 14(8.69-19.17) | **0.004** | 127 | 15(12.27-33.00) | **<0.0001** |
| Yes | 62 | 8(6.07-9.93) |  | 46 | 8(5.60-10.94) |  | 108 | 8(7.00-10.50) |  |

**Figure 3.**
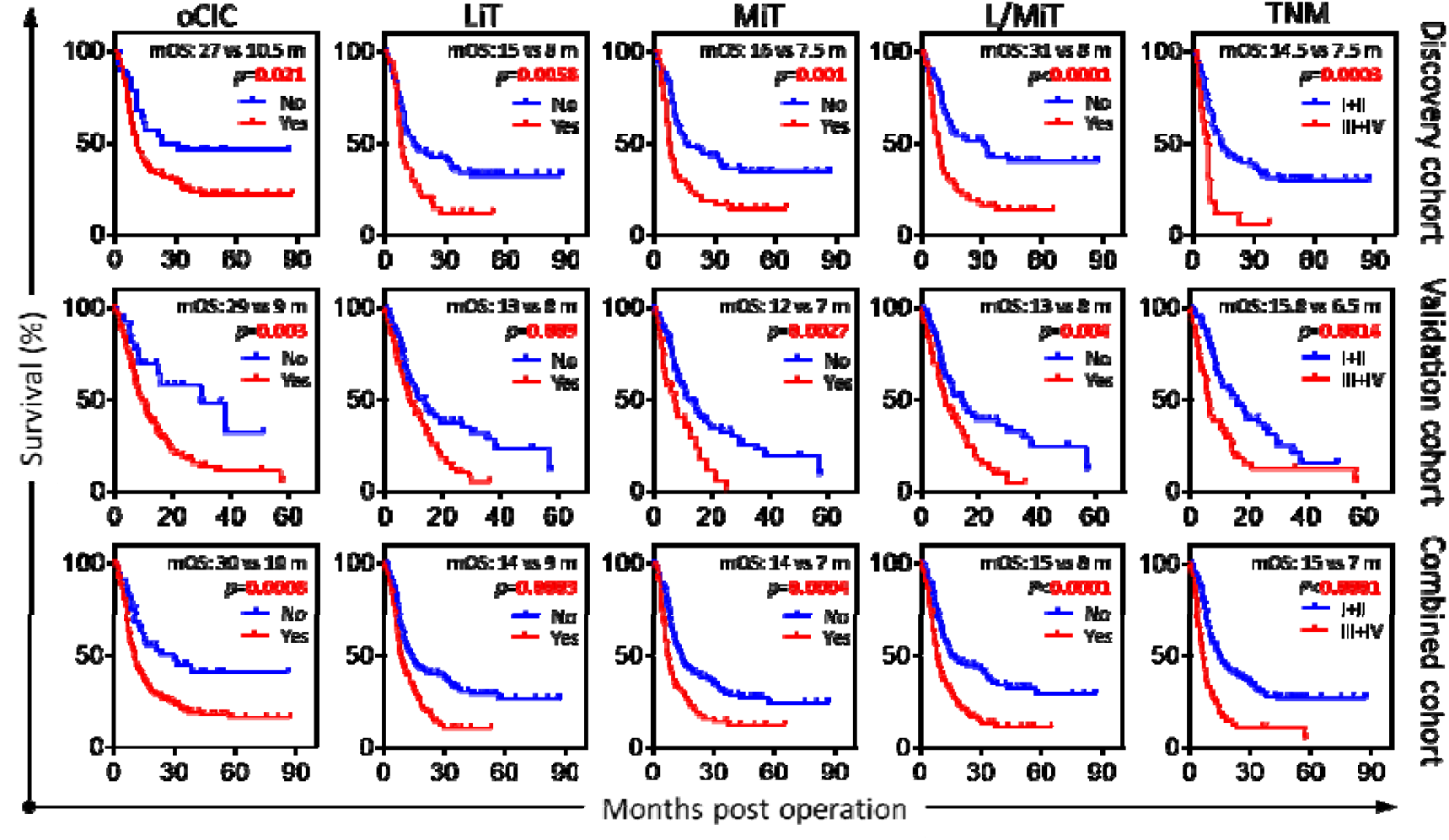
Cell-in-cell structures associated with overall survival of PDAC patients. Kaplan–Meier plotting of overall survival curves for indicated variables across different cohorts of patients.

### Heterotypic CICs are a prominent independent prognostic factor for PDAC

To analyze whether CICs and their subtypes could be independent prognostic factors for postoperative survival, we included all of the variables identified in univariate analysis (grade, TNM stage, LN invasion, distant metastasis, oCIC, TiT, LiM, MiT, and L/MiT) into multivariate survival analysis by using the Cox proportional hazards model. A stepwise selection process with these variables was performed. Unexpectedly, TNM stage was not an independent prognostic factor (HR = 1.642, 95%CI: 0.897–3.006, *p* = 0.108) in the discovery cohort, whereas L/MiT was the strongest risk factor for a poor prognosis, with a death hazard ratio of 1.850 (95%CI: 1.175–2.915, *p* = 0.008) (Table 3). Notably, the heterotypic L/MiT was the only variable that was consistently identified as an independent prognostic factor across the discovery, validation, and combined cohorts of patients (Table 3), suggesting that L/MiT could be a dominant contributor to a poor prognosis.

**Table 3.** Multivariate analysis of prognostic parameters for PDAC.

| Prognostic parameters | Discovery cohort |  | Validation cohort |  | Combined cohort |  |
| --- | --- | --- | --- | --- | --- | --- |
|  | HR(95%CI) | P | HR(95%CI) | P | HR(95%CI) | P |
| **Grade**<br>(I+II vs. III) | **1.775**<br>(1.133-2.781) | **0.0122** |  |  | **1.414**<br>(1.001-1.998) | **0.0493** |
| **N stage**<br>(Yes vs. No) | **1.598**<br>(1.034-2.472) | **0.0349** |  |  | **1.266**<br>(0.927-1.730) | 0.1374 |
| **M stage**<br>(Yes vs. No) |  |  | **1.663**<br>(0.898-3.080) | 0.1056 |  |  |
| **TNM stage**<br>(I+II vs. III+IV) | **1.642**<br>(0.897-3.006) | 0.1080 | **1.658**<br>(1.023-2.688) | **0.0403** | **2.008**<br>(1.430-2.820) | **0.0001** |
| **L/MiT**<br>(Yes vs. No) | **1.850**<br>(1.175-2.915) | **0.0080** | **1.714**<br>(1.086-2.705) | **0.0207** | **1.738**<br>(1.276-2.369) | **0.0005** |

To directly evaluate L/MiT’s contribution to prognosis, we constructed a nomogram that incorporated all 5 independent prognostic factors identified (grade, N stage, M stage, TNM, and L/MiT). In the nomogram, each variable was assigned a score on a point scale based on its predictive power according to our multivariate analysis. As shown in Figure 4, despite the fluctuations in the performance of the traditional variables across the discovery, validation, and combined cohorts of patients, L/MiT consistently dominated over all the traditional factors in predicting patient survival (Figure 4A–C). By locating the total score from all variables on the total point scale, the probability of patient survival could be estimated at a specified time point (14.0 months). As shown in Figure 4D–F, incorporating L/MiT improved the prediction performance (AUC from 0.710 to 0.767 for the discovery cohort, 0.658 to 0.674 for the validation cohort, and 0.661 to 0.696 for the combined cohort) at the time point of 14.0 months. We also calculated the AUC values at earlier survival time points including Q1 (6 months) and median (10.6 months), which reported similar results as shown in Table S8.

**Figure 4.**
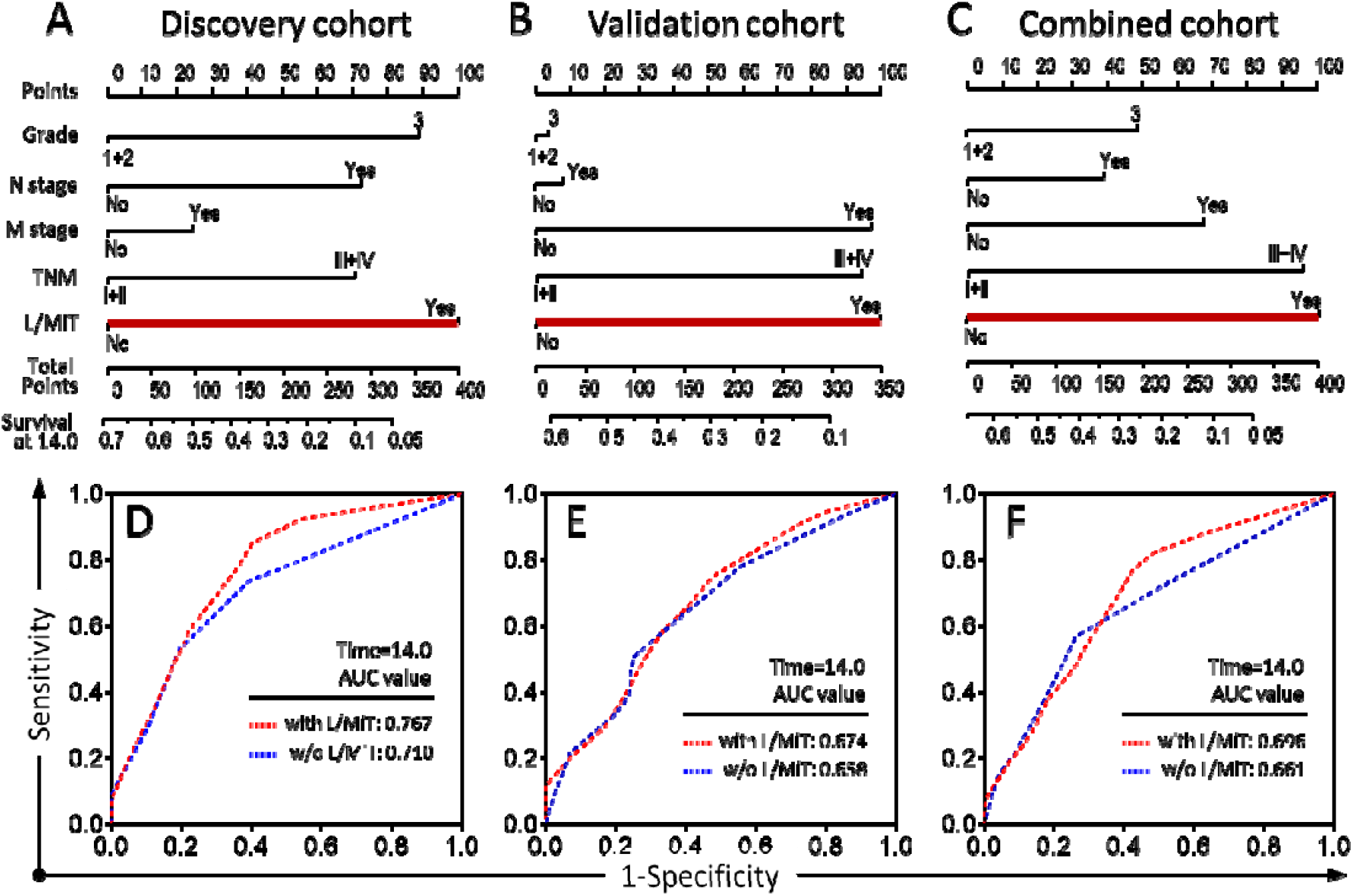
Contribution of heterotypic L/MiT to survival prediction by nomogram analysis. (**A–C**) Nomogram analysis with 5 independent prognostic factors identified (histological grade, N, M, and TNM stage, and L/MiT). L/MiT stands out as the dominant prognostic contributor across the discovery, validation, and combined cohorts of patients. (**D–F**) The AUC calculation for nomogram analysis of different cohorts of patients in the presence or absence of L/MiT.

### Heterotypic CICs preferentially impact the survival of young female patients with resectable PDAC

Pearson’s chi-squared test indicated that heterotypic CICs were associated with a late TNM stage and a higher histological grade (Table S1), which suggests that they may not be able to independently predict outcomes for late-PDAC patients. Instead, heterotypic L/MiT may, in specific cases, be an independent prognostic factor for resectable PDAC. To test this hypothesis, we stratified the combined cohort of patients by TNM stage (I+II vs. III+IV) or histological grade (1+2 vs. 3) for multivariate survival analysis. Traditional variables identified in univariate analysis (grade, N, M, and TNM stage) except for stratifiers, and all CICs (oCICs, TiT, LiT, MiT, TiM, and L/MiT), were examined by Cox proportional hazards model. As shown in Table 4, L/MiT was the only prognostic factor that could independently predict postoperative OS, specifically in patients of an early TNM stage, and the death hazard ratio increased (HR = 2.369, 95%:1.642–3.418; *p* < 0.0001) in comparison with that of the unstratified cohort (HR = 1.738, 95%:1.276–2.369; *p* = 0.0005) (Table 3). The selectivity of L/MiT was also applied to grade-stratified patients while TNM stage demonstrated a similar impact on patients of both low (1+2) and high (3) histological grade (Table 5). Further analysis indicated that L/MiT may specifically impact patients of TNM II (Table S3) and grade 2 (Table S4). Moreover, exhaustive analysis identified L/MiT as a highly selective predictor of a poor outcome in young (HR = 11.609, 95%: 3.872–34.81; *p* < 0.0001) (Table 6) and female (HR = 9.546, 95%: 2.566–35.52; *p* = 0.0008) (Table 7) patients while demonstrating no preference for tumor site, N stage, or M stage (Table S5–S7). The nomogram construction and AUC analysis confirmed that at 14 months, L/MiT was a selective prognostic classifier that predicts decreased survival in young female patients with resectable PDAC (Figure 5).

**Table 4.** Multivariate analysis of prognostic parameters for PDAC stratified by TNM2.

| Prognostic parameters | TNM1/2 (n = 175) |  | TNM3/4 (n = 60) |  |
| --- | --- | --- | --- | --- |
|  | HR(95%CI) | P | HR(95%CI) | P |
| **L/MiT** (Yes vs. No) | **2.3692**(1.642-3.418) | **<0.0001** |  |  |

**Table 5.**
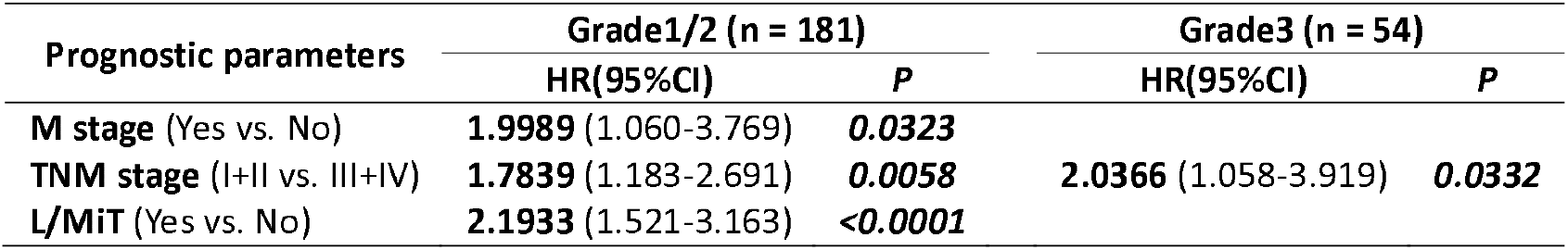
Multivariate analysis of prognostic parameters for PDAC stratified by Grade2.

**Table 6.** Multivariate analysis of prognostic parameters for PDAC stratified by age.

| Prognostic parameters | <=60 (n = 113) |  | >60 (n = 122) |  |
| --- | --- | --- | --- | --- |
|  | HR(95%CI) | P | HR(95%CI) | P |
| **Grade** (I+II vs. III) |  |  | **2.2319** (1.339-3.721) | **0.0021** |
| **N stage** (Yes vs. No) | **1.7430** (1.106-2.748) | **0.0167** |  |  |
| **TNM stage** (I+II vs. III+IV) | **2.6303** (1.527-4.531) | **0.0005** | **2.3120** (1.460-3.661) | **0.0004** |
| **TiT** (Yes vs. No) |  |  | **2.0167** (1.101-3.694) | **0.0231** |
| **TiM** (Yes vs. No) |  |  | **2.0935** (1.149-3.812) | **0.0157** |
| **L/MiT** (Yes vs. No) | **11.6097**(3.872-34.81) | **<0.0001** |  |  |

**Table 7.**
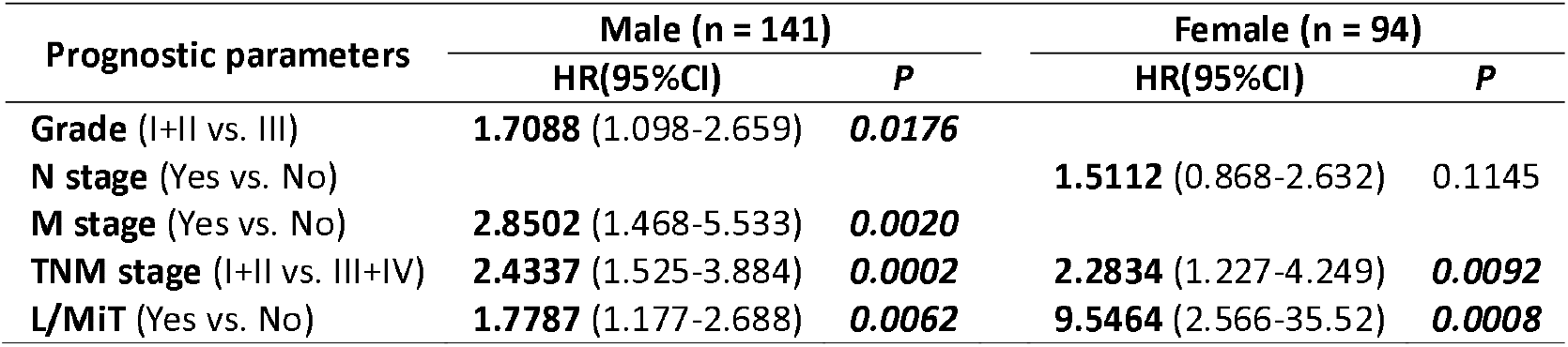
Multivariate analysis of prognostic parameters for PDAC stratified by sex.

| Prognostic parameters | Male (n = 141) |  | Female (n = 94) |  |
| --- | --- | --- | --- | --- |
|  | HR(95%CI) | P | HR(95%CI) | P |
| **Grade** (I+II vs. III) | **1.7088** (1.098-2.659) | **0.0176** |  |  |
| **N stage** (Yes vs. No) |  |  | **1.5112** (0.868-2.632) | 0.1145 |
| **M stage** (Yes vs. No) | **2.8502** (1.468-5.533) | **0.0020** |  |  |
| **TNM stage** (I+II vs. III+IV) | **2.4337** (1.525-3.884) | **0.0002** | **2.2834** (1.227-4.249) | **0.0092** |
| **L/MiT** (Yes vs. No) | **1.7787** (1.177-2.688) | **0.0062** | **9.5464** (2.566-35.52) | **0.0008** |

**Figure 5.**
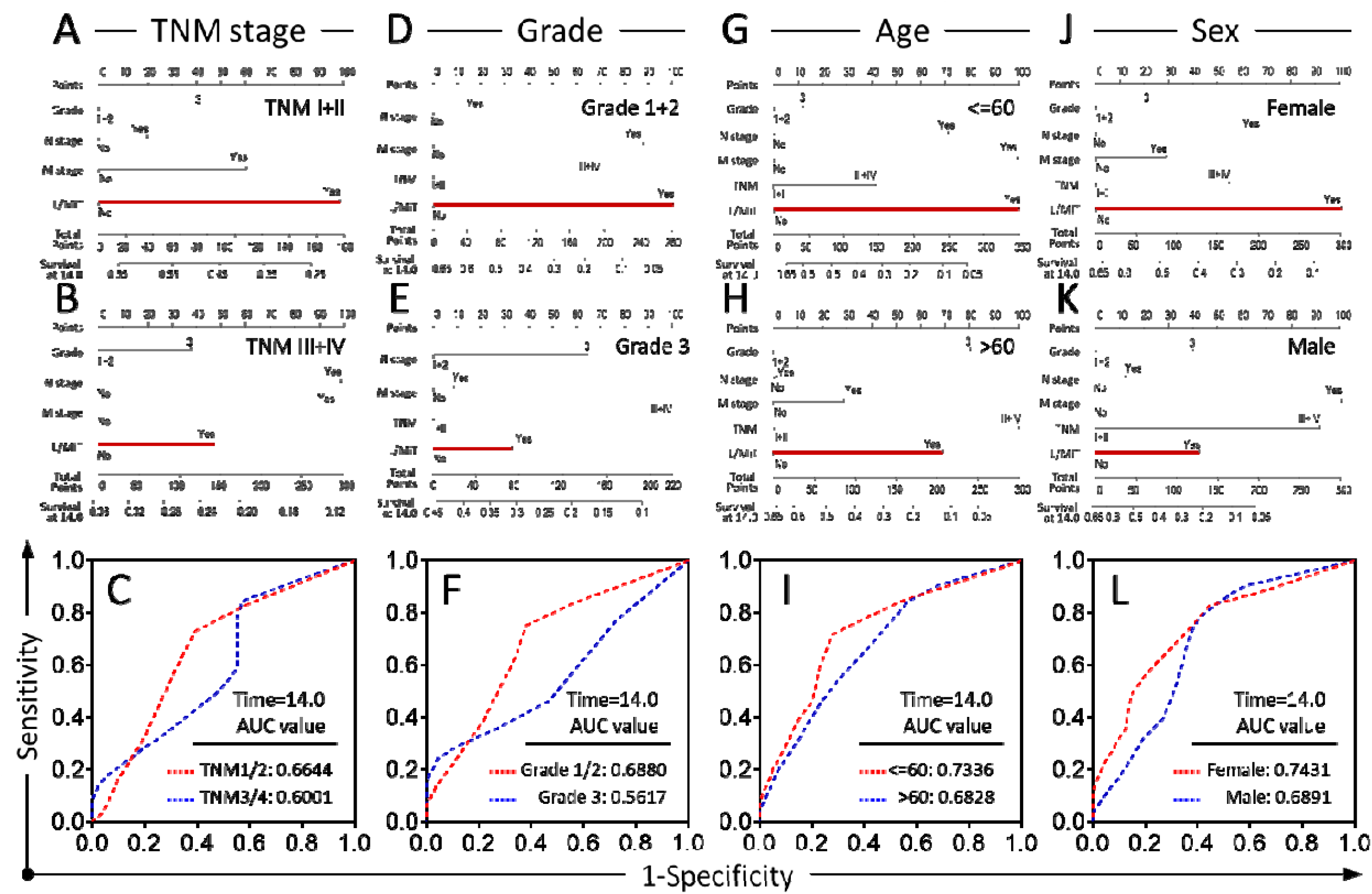
L/MiT selectively impacts overall survival of certain groups of PDAC patients. Nomogram and AUC analysis in combined cohort of patients stratified by TNM stage (I+II vs. III+IV) (**A–C**), histological grade (1+2 vs. 3) (**D–F**), age (≤ 60 vs. > 60) (**G–I**), and sex (female vs male) (**J–L**). L/MiT plays dominant role in young female patients with resectable PDAC.

## Discussion

Overall, our data support the hypothesis that subtyped CICs are promising prognostic markers for human PDAC, and that the presence of CICs is generally associated with a shorter survival time. Meanwhile, the prognostic performance of oCICs was profoundly affected by its subtype composition with the heterotypic CIC being a predominant factor. Furthermore, the active interactions between different CIC subtypes in patient prognosis were consistent with their intrinsically different biological functions. First, the presence of MiT or LiT may indicate that there is an even more malignant phenotype of tumor cell compared to TiT, as they (MiT and LiT) render tumor cells to kill macrophages, T cells, and/or natural killer cells that are designed to kill tumor cells, resulting in a form of immune evasion ^23^. Consistent with this notion, the presence of either LiT, MiT, or L/MiT was significantly associated with shorter patient OS, which also reinforces the concept that compromised immunity drives PDAC progression ^9^. Second, the presence of TiM may indicate the occurrence of immune activation, whereby tumor cells are eliminated by macrophage phagocytosis which has been shown to be a potential tumor therapy ^29, 30^. In agreement with this notion, a 65-year-old male patient, whose tumor tissue had a high level of TiM (12/core), survived for as long as 79 months as of his last visit, despite being diagnosed as histological grade 3. Our findings may also help to explain the unexpected protective role of CICs in PDAC metastasis, as reported in a study by Cano *et al*. ^25^ in which CD68^+^ CICs were identified but were not quantitatively subtyped. On these grounds, we can speculate that perhaps a considerable presence of TiM actively prevents metastasis. Intriguingly, TiT by entosis, a non-apoptotic cell death mechanism ^21^, was recently shown to be a mechanism of cell competition which promotes selection of malignant clones ^31-33^. The data here therefore suggests that a similar competitive mechanism may work in heterotypic CICs, resulting in tumors evolving to become more immune-resistant ^33^, and, as such, further functional validation is warranted.

Marker selection is an important part of CIC subtyping. Based on our preliminary study, E-cadherin, CD68, and CD45 are ideal for subtyping PDAC. This study, in line with previous research ^34^, found that, in comparison with adjacent non-cancerous tissues, downregulation of E-cadherin expression was common in the PDAC tissues. However, only 3 tissues saw a complete loss of E-cadherin expression (Figure S1F). In these cases, the cell morphology and CICs were identified by overexposure of background fluorescence assisted by H&E staining. Meanwhile, although CD163 was another accepted marker for macrophage, it was not a good marker for labeling macrophages participating in CIC formation ^16^. Instead, CD68 turned out to work well as an identifier of CICs in breast cancer ^35-37^, esophageal cancer (unpublished data), and pancreatic cancer as illustrated in this study.

An interesting finding of this work is that heterotypic L/MiT preferentially impacts certain groups of patients, specifically young women with early-stage PDAC (TNM I+II, or grade 1+2) (Table 4–7). In short, the presence of L/MiT in resectable PDAC tissues could independently predict poor outcomes for young female patients, whereas the survival of those without L/MiT was substantially longer. Though the underlying mechanisms warrant further investigation, this selective impact may reflect a dynamic role of a specified mechanism in different contexts. It is widely accepted that the development and progression of cancer is a net outcome of balance between driver and blocker factors ^38^. Each factor may dominate cancer progression in a defined context. We assumed that young and/or female PDACs were primarily promoted by the factors of active drivers, where simultaneous loss of blockers, such as immune surveillance by killing immune cells via L/MiT, would significantly potentiate cancer progression and predict poor prognosis. Therefore, our finding is not only informative and helpful for clinical practice but also provocative for further mechanistic investigations. It is conceivable that the formation of heterotypic L/MiT may surrogate the occurrence of specific oncogenic mutations, which, despite being invisible to traditional pathology in resectable PDAC, confer cancer cells the ability to cannibalize immune cells. Hence, exploring the molecular mechanisms underlying heterotypic CIC formation may help identify novel therapeutic targets for resectable PDAC with L/MiT and may benefit patient survival.

One of the most important implications of this work is the application of CICs as a functional index for patient diagnosis and prognosis. Current histological diagnosis largely depends on traditional pathology or molecular pathology, which generally produce information focusing on individual cells or molecules. Because tumors are quite heterogeneous, both in terms of morphology and genetics ^39-41^, multiple histological parameters are generally required for a relative improvement in tumor malignancy and patient prognosis predictions. Therefore, simple functional parameters that could readout tumor malignancy would be favored by clinicians. CICs arise from active cell-cell interactions between different types of cells within heterogeneous tumor microenvironments ^16, 17^. Formation of CICs generally leads to different functional outcomes of inner and outer cells, which can promote the growth of the outer cells while killing the inner cells ^42^. During CIC formation, the identities of inner or outer cells are genetically regulated ^36^, and oncogenic mutations such as *Kras* V12 are able to confer tumor cells outer/winner identity by inhibiting actomyosin contraction ^31^ while genetic inactivation of tumor suppressor CDKN2a or activation of p53 signaling leads cells to be internalized as inner/loser ^36, 43^. Thus, CIC is an ideal functional candidate to identify complex intercellular interactions and complicated intracellular signaling crosstalk. Consistent with this notion, our data in this work demonstrated that both oCICs and subtyped CICs (TiT, MiT, L/MiT) were able to predict patient prognoses with a performance comparable or even superior to traditional parameters such as TNM staging. Moreover, CICs were found to be a useful diagnostic indicator to differentiate benignity from malignancy in urothelial carcinoma, malignant mesothelioma, and effusion/urine cytology ^44-49^. And a high number of CICs were identified as an adverse prognostic factor for overall survival of patients with head and neck cancers ^27^, while in early breast cancer, CICs were found capable of selectively impacting patient survival in different categories and significantly contributing to the prediction of patient outcomes ^35^. Accordingly, we propose CIC profiling as a promising method to assist with tumor diagnosis and patient prognosis. It may constitute an essential part of an emerging functional pathology that will improve the performance of both traditional and molecular pathology.

According to our findings above, PDAC patients with CIC-positive samples may be at a higher risk of succumbing to poor survival even though they are pathologically at a low histological grade and early TNM stage. For such cases, active treatments and more frequent follow-up should be adopted. It should be noted that our data suggests that L/MiT preferentially impacted young and female patients, particularly those at early-stage, but the predictive values of other CICs subtypes may not be underscored across the entire patient cohort considering the limitations of this study, which is discussed below. Therefore, we recommend that CIC profiling should be performed post-operatively for all PDAC patients together with traditional pathology by using the paraffin-imbedded tumor sections. The consequences of the study reported here, which could be extended to other cancer types, are two-fold, namely: a) the study provides a readout to assess immune evasion and predict prognosis in PDAC; b) the study also supports the notion that the presence of L/MiT associates with cancer progression and shorter survival in cancer patients.

Despite these implications, the impact of the present study was limited by several factors. First, the retrospective nature of this study needs further validation through a prospective study. Second, although this is currently the largest subtype-based CIC profiling of human cancers, the tissue sample size should be expanded in future studies for further confirmation. Third, since commercial TMA was used to explore the prognostic value of CICs in this study, some information, such as neoadjuvant chemotherapy, and subsequent treatments following surgery and date of first relapse, were not available; this prevents us from evaluating the effects various treatments have on both patient survival and disease-free survival (DFS). These considerations warrant further investigation in the future.

In summary, this study reported the first subtype-based CIC profiling in human PDAC, and identified oCICs and its heterotypic subtypes (LiT, TiM, and L/MiT) as valuable prognostic markers in predicting patient survival in a specified group. L/MiT was identified as a potent adverse prognostic marker impacting young female patients with early-stage PDAC. Our work also supports functional pathology with CIC profiling as a novel input for traditional pathology, and the promise it holds for improving clinical diagnosis and guiding cancer therapy.

## Data Availability

The raw/processed data required to reproduce these findings cannot be shared at this time as the data also forms part of an ongoing study.

## Acknowledgements

The authors appreciate the academic support from the AME Pancreatic Cancer Collaborative Group, and professional advice from Dr. Ryan M. Thomas (University of Florida, USA), Dr. Roberta Elisa Rossi (Università degli Studi di Milano, Italy), Dr. Paul Zarogoulidis (“Bioclinic” Private Hospital, Thessaloniki, Greece), Dr. Benedetto Ielpo (University Hospital of Leon, Spain), Dr. Nicolas Jonckheere (Univ Lille, CNRS, Inserm, France), Dr. Marco Falasca (Curtin University, Australia), and Dr. Faustino Mollinedo (Consejo Superior de Investigaciones Científicas, Spain).

---

**Figure S1.**
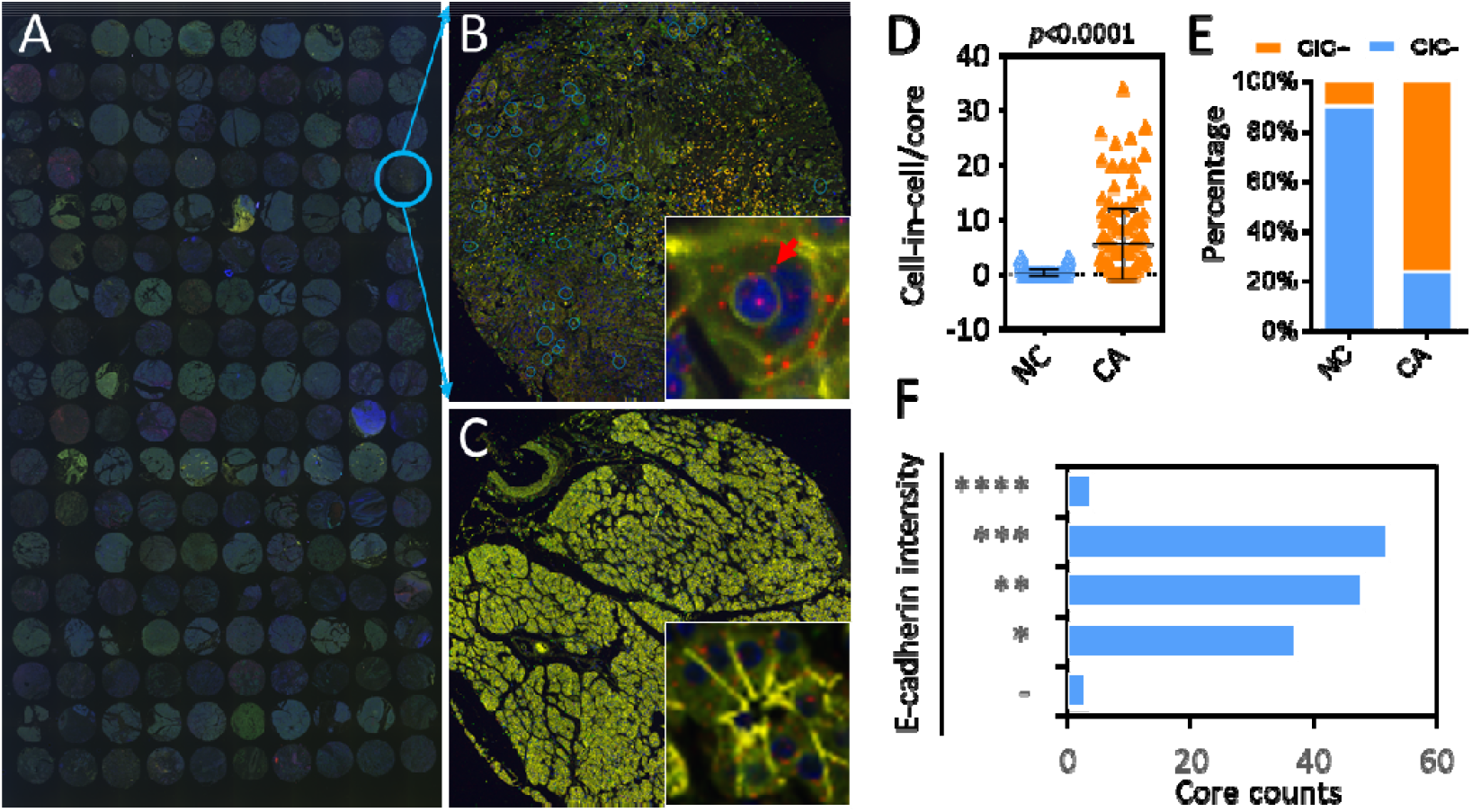
Detecting CIC structures in tissue microarray (TMA) of human PDAC. (A–C) Composite images of the whole TMA slide (A), a single representative cancer tissue core (B), and a non-malignant tissue core (C). Inserted images show a typical CIC structure or acinar. The arrow indicates the inner cell. E-cadherin staining (in yellow) indicates intercellular contact. (D) The profiles of all CICs detected are depicted as CIC number per core in cancer (CA) and non-cancerous (NC) tissues. (E) The CIC compositions of cancer (CA) and non-cancerous (NC) tissues. (**F**) E–cadherin expression profile in pancreatic cancerous tissue cores. “–” for negative, “****” for strongest expression comparable to the adjacent non-malignant tissues. n = 144.

**Figure S2.**
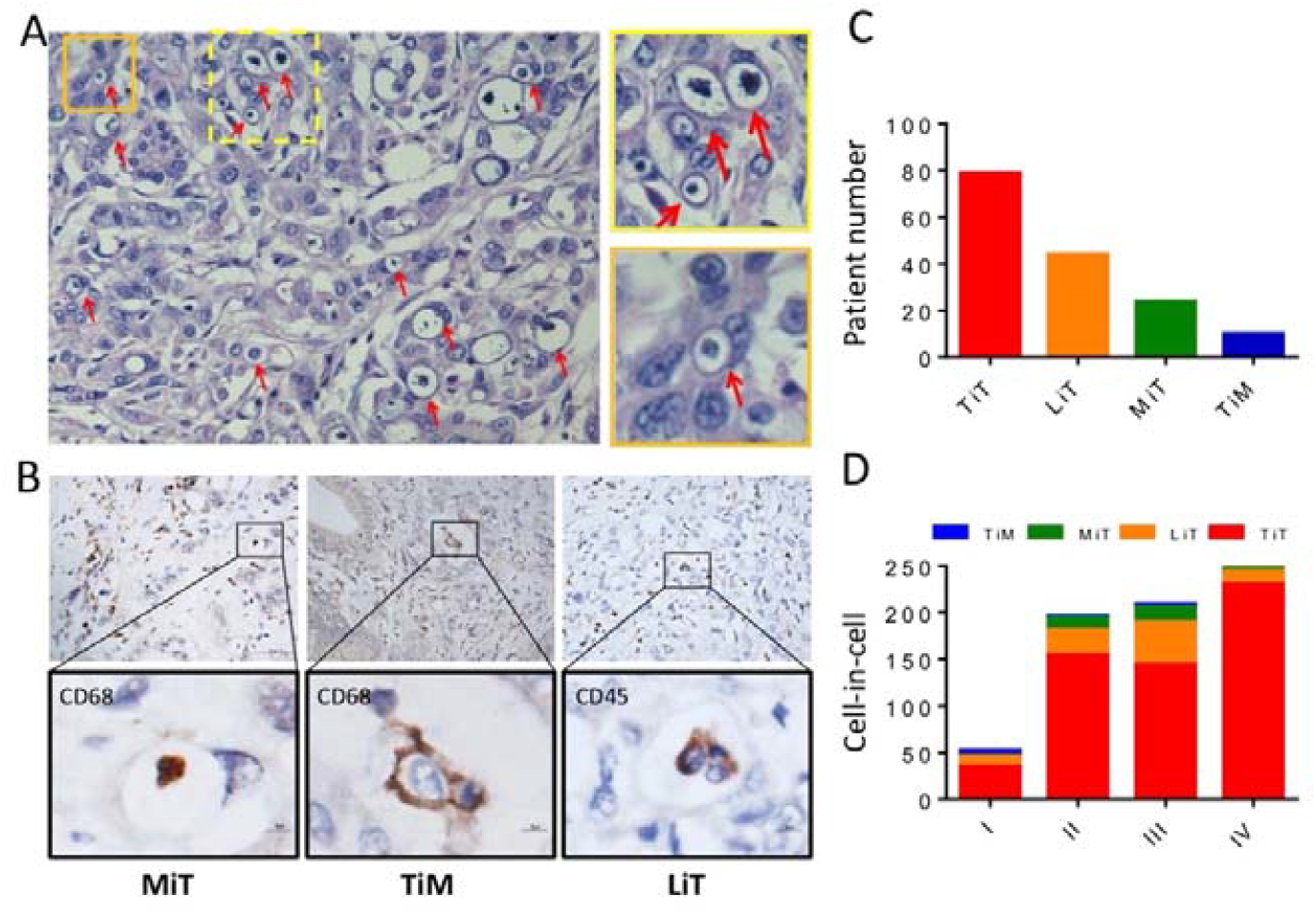
Detecting CIC structures in human PDAC of the validation cohort. (A) CIC structures indicated by arrows in PDAC tissues stained by H&E. Boxed regions are zoomed in at the right. (B) CIC structures in PDAC tissues stained with antibodies for CD68 or CD45, respectively, by IHC. Boxed regions are zoomed in at the bottom. (C) Number of tissues positive in each CIC subtype. (D) The compositions of CIC subtypes in different TNM stages.

**Figure S3.**
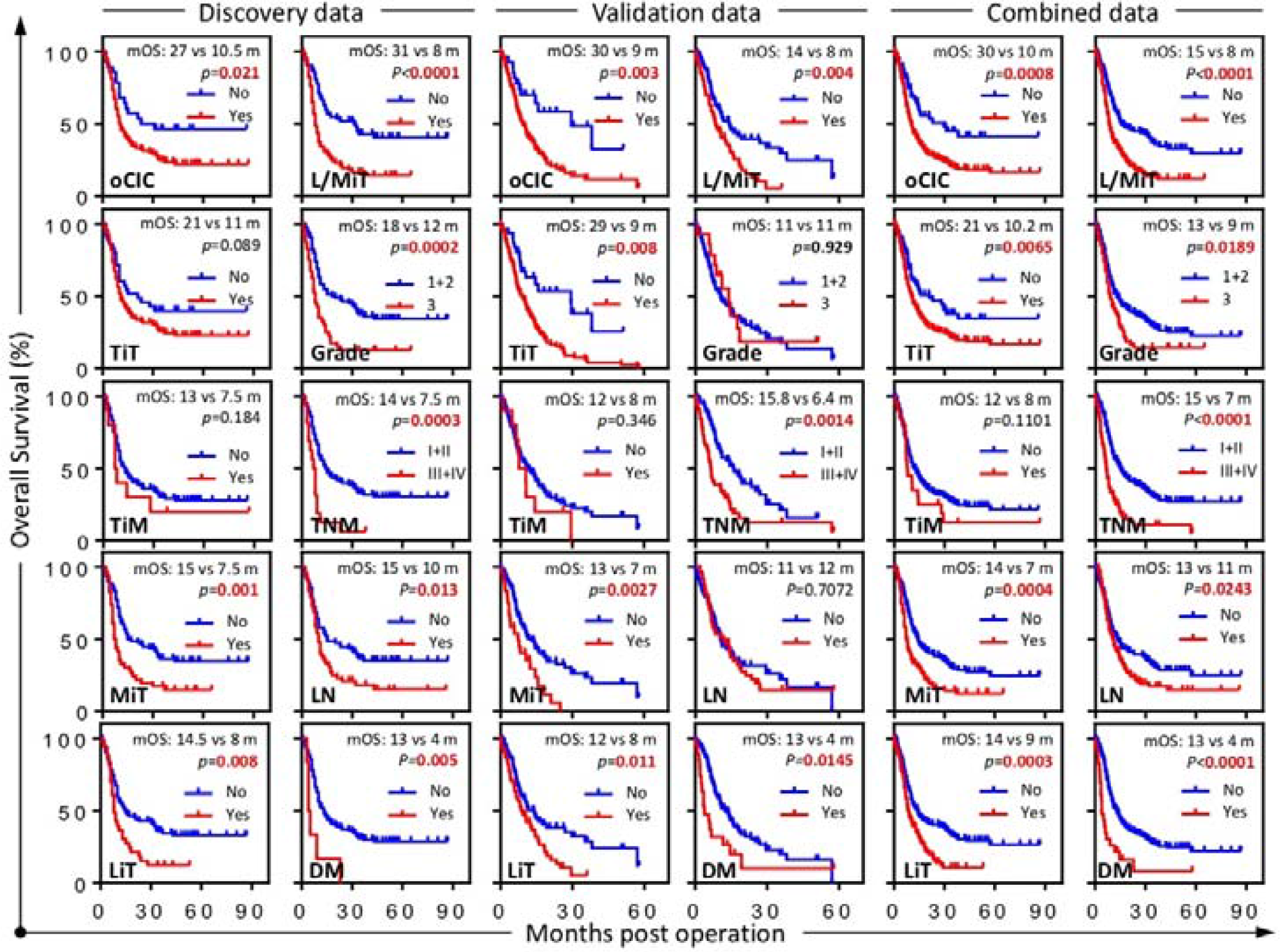
Kaplan–Meier plotting of overall survival curves for indicated variables across the different cohorts of patients. LN for lymph node invasion; DM for distant metastasis.

**Table S1a:**
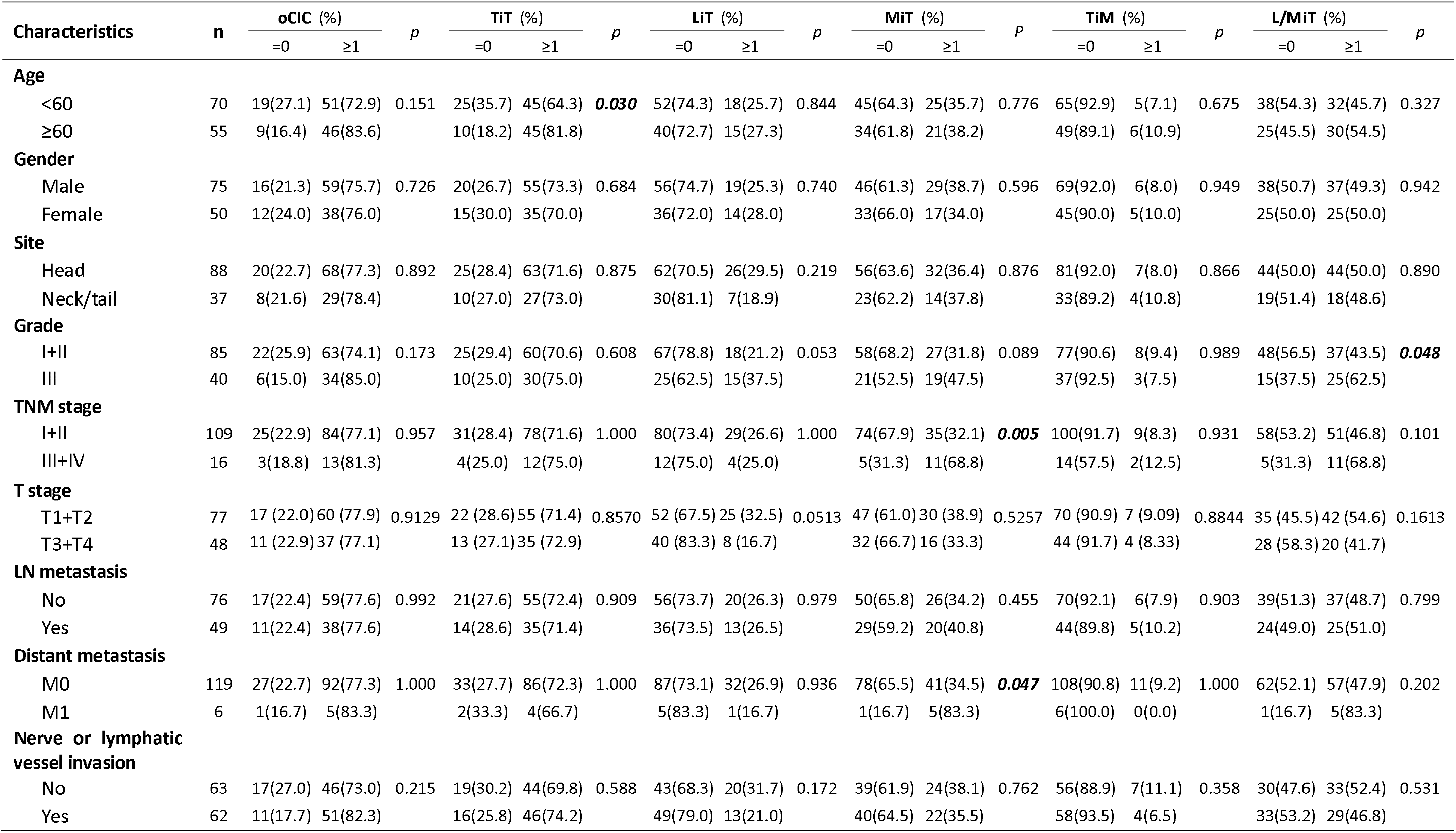
Association of CIC subtypes with clinicopathological characteristics for the discovery dataset.

**Table S1b:**
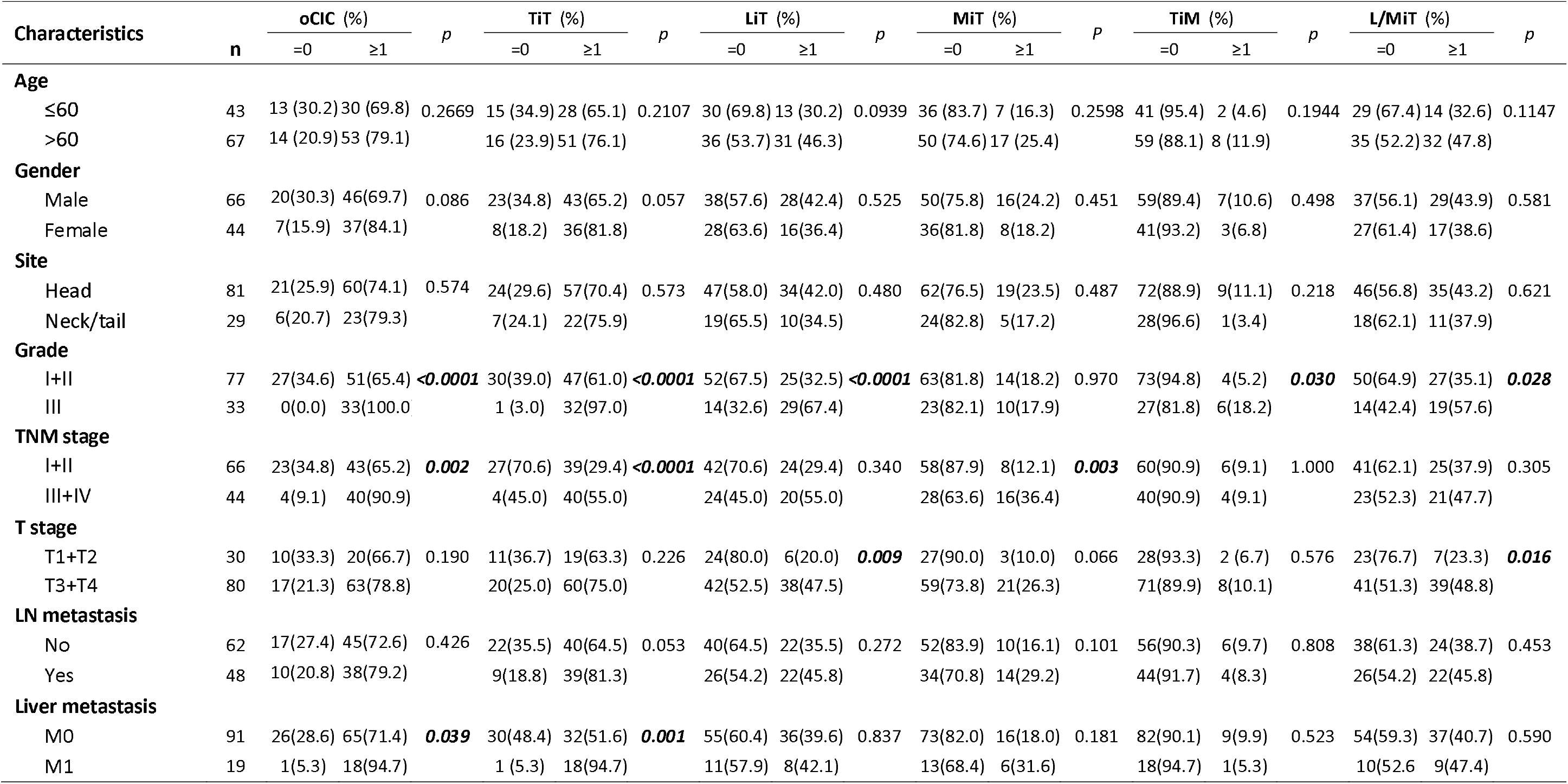
Association of CIC subtypes with clinicopathological characteristics for the validation dataset.

**Table 1c:**
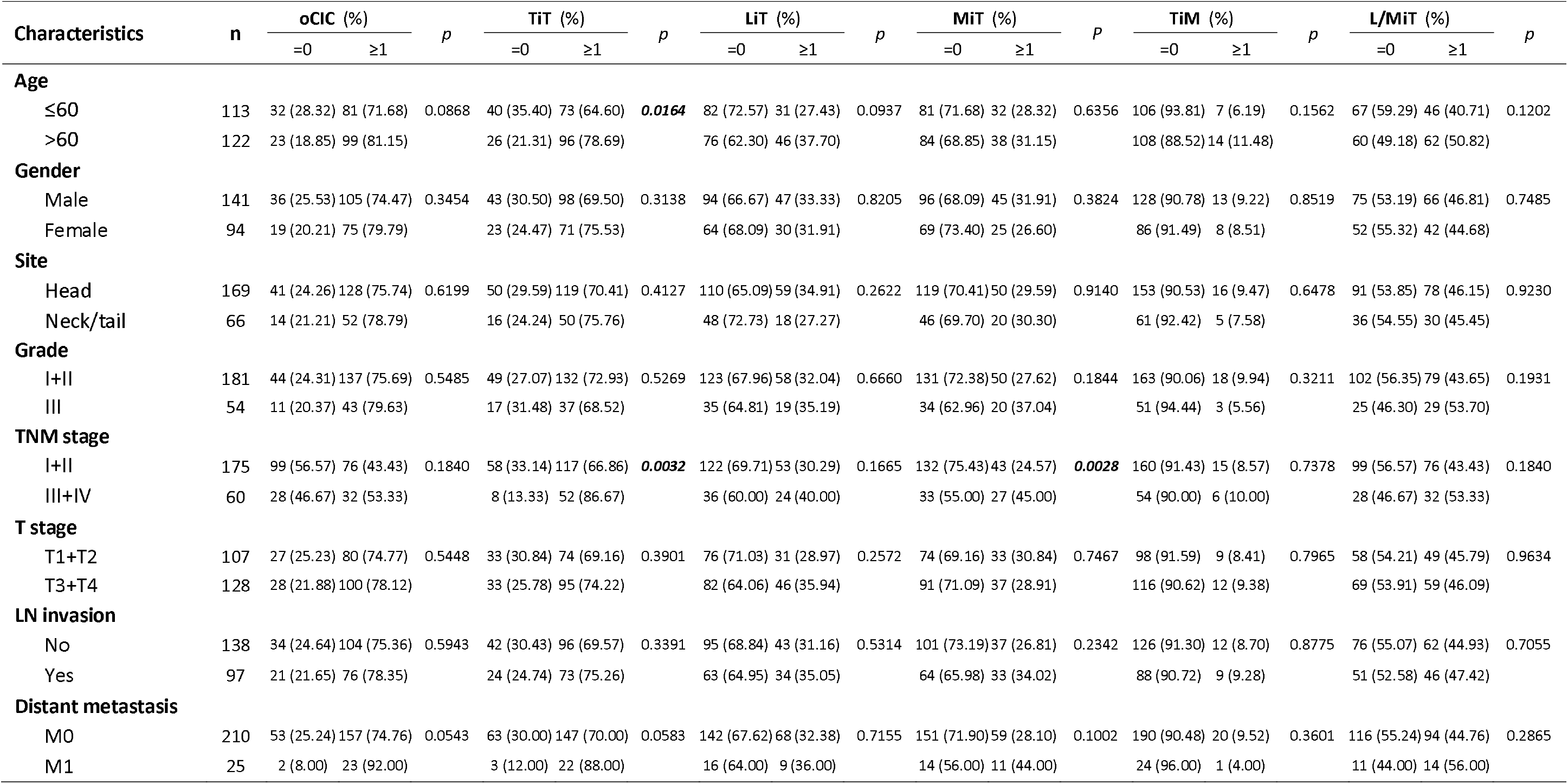
Association of CIC subtypes with clinicopathological characteristics for the combined dataset.

**Table S2.** Univariate analysis of prognostic parameters for survival in PDAC patients by Cox regression analysis.

| Characteristics | n (125) | Discovery cohort | p(COX) | n (110) | Validation cohort | p(COX) | n(235) | Combined cohort | p(COX) |
| --- | --- | --- | --- | --- | --- | --- | --- | --- | --- |
|  |  | HR (95% CI) |  |  | HR (95% CI) |  |  | HR (95% CI) |  |
| **Age** (≥60/<60) | 55/70 | 0.956(0.629-1.453) | 0.832 | 67/43 | 0.976(0.586-1.516) | 0.807 | 122/113 | 1.020 (0.753-1.383) | 0.8962 |
| **Sex** (Female/Male) | 50/75 | 0.764(0.497-1.174) | 0.219 | 44/66 | 1.070(0.682-1.679) | 0.768 | 94/141 | 0.902(0.662-1.229) | 0.5135 |
| **Site** (Body+tail/Head) | 37/88 | 0.875(0.555-1.381) | 0.567 | 29/81 | 0.726(0.419-1.258) | 0.253 | 66/169 | 0.801(0.565-1.135) | 0.2122 |
| **Grade** (III/I+II) | 40/85 | 2.215(1.403-3.311) | **<0.0001** | 33/77 | 1.022 (0.637-1.640) | 0.929 | 54/181 | 2.190(1.426-3.364) | **0.0003** |
| **TNM stage** (III+IV/I+II) | 16/109 | 2.629(1.491-4.634) | **0.001** | 44/66 | 2.029(1.302-3.160) | **0.002** | 60/175 | 2.281(1.601-3.250) | **<0.0001** |
| **T stage** (T3+T4/T1+T2) | 48/77 | 0.738(0.476-1.145) | 0.169 | 80/30 | 1.572(0.954-2.589) | 0.076 | 128/107 | 1.032(0.757-1.407) | 0.8417 |
| **LN invasion** (Yes/No) | 49/76 | 1.673(1.101-2.542) | **0.016** | 48/62 | 1.088(0.701-1.688) | 0.707 | 97/138 | 1.687 (1.110-2.564) | **0.0143** |
| **Distant metastasis** (M1 /M0) | 6/119 | 3.075(1.329-7.114) | **0.009** | 19/91 | 1.975(1.133-3.441) | **0.016** | 25/210 | 3.172 (1.372-7.336) | **0.007** |
| **oClCs** (Yes/No) | 97/28 | 1.874(1.075-3.266) | **0.027** | 83/27 | 2.431(1.313-4.499) | **0.005** | 180/55 | 1.981(1.320-2.972) | **0.001** |
| **TiT** (Yes/No) | 90/35 | 1.509(0.925-2.462) | 0.099 | 79/31 | 2.063(1.190-3.573) | **0.01** | 169/66 | 1.648(1.149-2.365) | **0.0067** |
| **LiT** (Yes/No) | 33/92 | 1.798(1.149-2.812) | **0.010** | 44/66 | 1.771(1.134-2.767) | **0.012** | 77/158 | 1.701(1.240-2.333) | **0.001** |
| **MiT** (Yes/No) | 46/79 | 1.932(1.268-2.943) | **0.002** | 24/86 | 2.140(1.287-3.560) | **0.003** | 70/165 | 1.971(1.294-3.002) | **0.0016** |
| **TiM** (Yes/No) | 11/114 | 1.574(0.79-3.137) | 0.198 | 10/100 | 1.395(0.695-2.799) | 0.349 | 21/214 | 1.463(0.897-2.388) | 0.1275 |
| **L/MiT** (Yes/No) | 62/63 | 2.249(1.470-3.441) | **<0.0001** | 46/64 | 1.907(1.220-2.981) | **0.005** | 108/127 | 1.984(1.464-2.688) | **<0.0001** |

**Table S3.** Multivariate analysis of prognostic parameters for PDAC stratified by TNM.

| Prognostic parameters | TNM1 (n = 65) |  | TNM2 (n = 110) |  | TNM3 (n = 35) |  | TNM4 (n = 25) |  |
| --- | --- | --- | --- | --- | --- | --- | --- | --- |
|  | HR(95%CI) | P | HR(95%CI) | P | HR(95%CI) | P | HR(95%CI) | P |
| **Grade**<br>(I+II vs. III) |  |  | **1.636**<br>(0.965-2.776) | 0.0678 |  |  |  |  |
| **MiT**<br>(Yes vs. No) |  |  | **1.704**<br>(0.906-3.205) | 0.0983 |  |  |  |  |
| **LiT**<br>(Yes vs. No) | **1.993**<br>(1.622-5.521) | **0.0005** |  |  | **0.4879**<br>(0.226-1.054) | 0.0679 |  |  |
| **L/MiT**<br>(Yes vs. No) |  |  | **1.904**<br>(1.048-3.461) | **0.0346** |  |  |  |  |

**Table S4.** Multivariate analysis of prognostic parameters for PDAC stratified by grade.

| Prognostic parameters | Grade1 (n = 36) |  | Grade2 (n = 145) |  | Grade3 (n = 54) |  |
| --- | --- | --- | --- | --- | --- | --- |
|  | HR(95%CI) | P | HR(95%CI) | P | HR(95%CI) | P |
| **N stage**<br>(Yes vs. No) | **0.3602**<br>(0.145-0.894) | **0.0276** | **1.4230**<br>(0.939-2.156) | **0.0863** |  |  |
| **M stage**<br>(Yes vs. No) | **3.7783**<br>(1.221-11.68) | **0.0210** | **2.3037**<br>(1.059-5.010) | **0.0353** |  |  |
| **TNM stage**<br>(I+II vs. III+IV) |  |  | **1.8931**<br>(1.165-3.076) | **0.0100** | **2.0366**<br>(1.058-3.919) | **0.0332** |
| **L/MiT**<br>(Yes vs. No) |  |  | **2.2716**<br>(1.517-3.402) | **0.0001** |  |  |

**Table S5.** Multivariate analysis of prognostic parameters for PDAC stratified by site.

| Prognostic parameters | Head (n = 169) |  | Body/tail (n = 66) |  |
| --- | --- | --- | --- | --- |
|  | HR(95%CI) | P | HR(95%CI) | P |
| **Grade** (I+II vs. III) | **1.3941** (0.937-2.075) | 0.1014 |  |  |
| **N stage** (Yes vs. No) |  |  | **2.5745** (1.334-4.968) | **0.0048** |
| **M stage** (Yes vs. No) |  |  | **3.4845** (1.531-7.926) | **0.0029** |
| **TNM stage** (I+II vs. III+IV) | **2.1329** (1.443-3.152) | **0.0001** |  |  |
| **TiM** (Yes vs. No) |  |  | **2.6783** (0.988-7.263) | 0.0529 |
| **L/MiT** (Yes vs. No) | **1.8276** (1.277-2.656) | **0.0010** | **2.5390** (1.285-5.017) | **0.0073** |

**Table S6.**
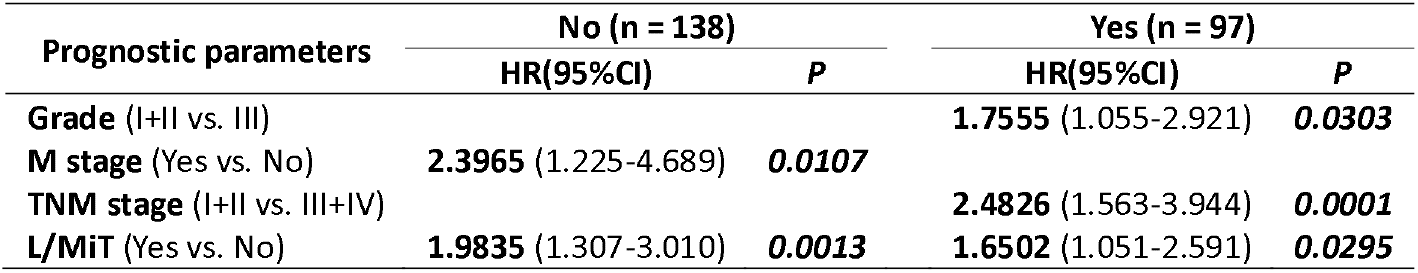
Multivariate analysis of prognostic parameters for PDAC stratified by N stage.

**Table S7.** Multivariate analysis of prognostic parameters for PDAC stratified by M stage.

| Prognostic parameters | No (n = 210) |  | Yes (n = 25) |  |
| --- | --- | --- | --- | --- |
|  | HR(95%CI) | P | HR(95%CI) | P |
| **Grade** (I+II vs. III) | **1.4794** (1.007-2.173) | **0.0461** | **0.2820** (0.066-1.211) | 0.0887 |
| **N stage** (Yes vs. No) | **1.3054** (1.225-4.689) | 0.1168 |  |  |
| **TNM stage** (I+II vs. III+IV) | **1.8578** (1.230-2.805) | **0.0032** | inf. (0.000-Inf) | 0.9979 |
| **L/MiT** (Yes vs. No) | **1.6514** (1.183-2.306) | **0.0032** | inf. (0.000-Inf) | 0.9995 |

**Table S8.** AUC values calculated at different survival time points by ROC analysis.

| Time points | 6 months |  | 10.6 months |  | 14 months |  |
| --- | --- | --- | --- | --- | --- | --- |
|  | with | w/o | with | w/o | with | w/o |
| **Discovery cohort** | 0.737 | 0.657 | 0.760 | 0.676 | 0.767 | 0.71 |
| **Validation cohort** | 0.725 | 0.719 | 0.672 | 0.668 | 0.674 | 0.658 |
| **Combined cohort** | 0.710 | 0.653 | 0.698 | 0.653 | 0.696 | 0.661 |

w/o: without

## Notes

**Financial support:** The authors declare that they have no competing financial interests. This work was supported by the National Key Research & Development Program of China (2016YFC1303303 to QS, 2018YFA0900804 to YZ, 2019YFA09003801 to QS), and the National Natural Science Foundation of China (81572799 to HH, 31671432 to QS, 31770975 to XW, 81972483 to MH).

### Competing Interest Statement

The authors have declared no competing interest.

### Funding Statement

This work was supported by the National Key Research & Development Program of China (2016YFC1303303 to QS, 2018YFA0900804 to YZ, 2019YFA09003801 to QS), and the National Natural Science Foundation of China (81572799 to HH, 31671432 to QS, 31770975 to XW, 81972483 to MH).

### Author Declarations

Human tissue microarrays (TMA) and "clinical-type" gene chips (CTGCs)were purchased from The Outdo Biotech. Tissue sections were collected from 110 resectable PDAC patients who had received surgery at the Department of Hepatobiliary Surgery, the First Affiliated Hospital of Sun Yat-sen University (Guangzhou, China), with the institutional research ethics committee reviewing and approving the research.

